# Climate effect on COVID-19 spread rate: an online surveillance tool

**DOI:** 10.1101/2020.03.26.20044727

**Authors:** Gil Caspi, Uri Shalit, Søren Lund Kristensen, Doron Aronson, Lilac Caspi, Oran Rossenberg, Avi Shina, Oren Caspi

## Abstract

**Background:** COVID-19 outbreak poses an unprecedented challenge for societies, healthcare organizations and economies. In the present analysis we coupled climate data with COVID-19 spread rates worldwide, and in a single country (USA).

**Methods:** Data of confirmed COVID-19 cases was derived from the COVID-19 Global Cases by the CSSE at Johns Hopkins University up to March 19, 2020. We assessed disease spread by two measures: replication rate (RR), the slope of the logarithmic curve of confirmed cases, and the rate of spread (RoS), the slope of the linear regression of the logarithmic curve.

**Results:** Based on predefined criteria, the mean COVID-19 RR was significantly lower in warm climate countries (0.12±0.02) compared with cold countries (0.24±0.01), (P<0.0001). Similarly, RoS was significantly lower in warm climate countries 0.12±0.02 vs. 0.25 ± 0.01 than in cold climate countries (P<0.001). In all countries (independent of climate classification) both RR and RoS displayed a moderate negative correlation with temperature R= -0.69, 95% confidence interval [CI], -0.87 to -0.36; P<0.001 and R= -0.72, 95% confidence interval [CI], -0.87 to -0.36; P<0.001, respectively. We identified a similar moderate negative correlation with the dew point temperature. Additional climate variables did not display a significant correlation with neither RR nor RoS. Finally, in an ancillary analysis, COVID-19 intra-country model using an inter-state analysis of the USA did not identify yet correlation between climate parameters and RR or RoS as of March, 19, 2020.

**Conclusions:** Our analysis suggests a plausible negative correlation between warmer climate and COVID-19 spread rate as defined by RR and RoS worldwide. This initial correlation should be interpreted cautiously and be further validated over time, the pandemic is at different stages in various countries as well as in regions within these countries. As such, some associations may be more affected by local transmission patterns rather than by climate. Importantly, we provide an online surveillance dashboard (https://covid19.net.technion.ac.il/) to further assess the association between climate parameters and outbreak dynamics worldwide as time goes by.

**Research in context:** *Evidence before this study:* The coronavirus, COVID-19 pandemic caused by the novel SARS-CoV 2, challenges healthcare organizations and economies worldwide. There have been previous reports describing the association between seasonal climactic variance and SARS-CoV 1 as well as the MERS infections, but the association with SARS-CoV 2 and climate has not been described extensively.

*Added value of this study:* Our analysis demonstrates a plausible negative correlation between warmer climate and COVID-19 spread rate as defined by RR and RoS worldwide in all countries with local transmission as of March 9, 2020. This initial correlation should be interpreted cautiously and be further validated over time. Importantly, we provide an online surveillance dashboard available at (https://covid19.net.technion.ac.il/) for further dynamic tracking of climate effect on COVID-19 disease spread rate worldwide and on intra-country analysis between USA states.

*Implications of all the available evidence:* Our findings of decreased replication and spread rates of COVID-19 in warm climates may suggest that the inevitable seasonal variance will alter the dynamic of the disease spread in both hemispheres in the coming months. However, we warrant a cautious interpretation of these findings given the fact that we are in the initial steps of this outbreak in many “warm” climate countries, the high variance of the data and the dynamic changes in the disease surveillance and the lack of correlation based on the limited data in the US. We hope that the online tool coupling COVID-19 data with climate data will assist in tracking the disease and tailoring the needed measures to contain it.

## Introduction

The coronavirus, COVID-19 pandemic, challenges healthcare organizations and economies worldwide. As of March 20, 2020, a total of 260,476 COVID-19 cases have been confirmed and 11,289 deaths. Transmission of COVID-19 is community-based unlike previous coronavirus outbreaks such as severe acute respiratory syndrome coronavirus (SARS-CoV) or the Middle East respiratory coronavirus (MERS-CoV) that were both mainly transmitted in the hospital setting^1,2^. Recent reports show that patients infected with COVID-19 are at high risk for severe morbidity (5% intensive care unit admissions) and mortality (1.4%)^3^, although highly dependent on age and prior comorbidity^4^. The reproduction number (R0), which defines the average number of cases directly generated by one case, for COVID-19 is estimated to be between 1.5-3.5^5,6^ and the reproduction efficacy may be influenced by cultural habits, population density and the country specific mitigation methods such as quarantine strategies as well as travel control measures^7-9^. An additional key factor that is of specific interest world-wide and a source for controversy is the effect of the climate on COVID-19 transmission^10^.

The spatio-temporal transmission of respiratory viruses such as influenza is highly associated with meteorological factors such as temperature, humidity and rainfall, with peak incidence occurring during winter^11^. The SARS-CoV epidemic in Beijing (2003) peaked during the early spring time and disease spread rate correlated with climate variables^12^. Similarly, SARS-CoV spread in Hong Kong (2003) was shown to negatively correlate with higher temperatures^13^.Unlike the SARS-CoV, the highest global seasonal occurrence of the MERS-CoV occurred during spring-summer periods in countries with warm climate such as Saudi-Arabia^14,15^ and high temperatures as well as low humidity were associated with increased disease spread rate^16^.

Given the variant vulnerability of different strains of coronavirus to climate, specifically temperature, there is a major global uncertainty regarding the continuing spread of COVID-19 during the coming months. Currently, the World Health Organization assumption is that COVID-19 spread will not be ameliorated during the summer period^17^. A surveillance tool assessing the correlation between the spread rate of COVID-19 and climate variables will be instrumental for societies, governments and health organizations worldwide. Analyzing the interaction between disease spread rate and climate may allow implementation of differential and precise mitigation measures for disease spread prevention, to tune healthcare routine ambulatory services and preparation strategies and to minimize the unnecessary dreadful impact of excessive quarantine strategies on psychosocial health and economies. Online dashboards have proven useful for global COVID-19 tracking. We have two aims in the current study. First, to compare the distinctive transmission efficacy of COVID-19 in countries with cold and warm climate based on the initial disease diagnosis trends worldwide. We consider this to be a cautious test case for examining the correlation between climate and COVID-19 disease spread. The second and more important aim, is to create an online up-to-date surveillance tool simultaneously presenting COVID-19 spread rate with relation to climate parameters.

## Methods

Data of confirmed COVID-19 cases was derived from the Coronavirus COVID-19 Global Cases by the Center for Systems Science and Engineering (CSSE) at Johns Hopkins University (JHU)^18^up to March 19, 2020. Countries with less than 50 diagnosed patients as well as countries not categorized as local transmission according to the WHO situation report as of March 9 were excluded in order to minimize confounding of imported disease transmission.

We prospectively defined “warm” and “cold” climate countries according to the following criteria: A “Cold” climate country was defined as a country with average temperatures below 15° Celsius degrees (<77° Fahrenheit) during the month of March10 and latitude line north of 40°. A “Warm” climate country was defined as a country with average March temperatures above 15° Celsius degrees (≥77° Fahrenheit). Based on the assumption that Italy has experienced an unproportionally high local disease spread not essentially affected by weather (in a similar manner to the Wuhan outbreak) we tested our model with and without the addition of Italy. We did not include China in our analysis given the unique circumstances associated with the country being the origin of the outbreak, the lag between outbreak and detection that may confound spread as well as the drastic mitigation steps applied. We assessed disease spread by two measures: The Replication Rate and the Rate of Spread.

Replication rate (RR) was defined as the slope of the logarithmic curve of the natural logarithm of the number of cases diagnosed in each country, starting from the day in which the total number of diagnosed cases was ≥30. The choice of 30 for the point we start counting diagnosed cases was chosen as a cut-off based on 2 standard deviations from the mean diagnosed patients in countries with imported cases only (based on WHO situation report at March 5, 2020) We calculated the slope of a sliding window of size (dT), where we chose dT=3. Let *C*_*t*_ be the number of validated cases of COVID-19 for each country at day t.

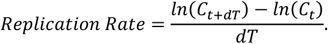

Rate of Spread (RoS) was calculated based on the method presented by Sajadi et al.^10^. It is calculated by running a linear regression of ln(Confirmed Cases) on time, and taking RoS to be the slope coefficient of the regression. We used a 7-day sliding window, as in Sajadi et al. ^10^.

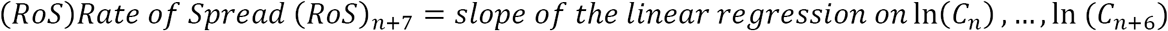

The calculation of the RoS was conducted by using a window for regression that does not include any missing values. From the RoS one can estimate the doubling time of cases: 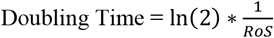.^19^

### Databases used

Country population data was taken from the United Nations website, the Department of Economic and Social Affairs Population (https://www.un.org/en/development/desa/population/index.asp).COVID-19 diagnostic test numbers were taken from https://ourworldindata.org/covid-testing updated for March, 20, 2020. Climate data was derived from www.weatherbase.com and based on country capital historical average climate for the month of March (based on weather data at and missing data was added from en.climate-data.org). Average temperature, precipitation in mm, morning and evening humidity, dew point (the temperature to which air must be cooled in order to reach saturation with water) and wind speed (km/h) were collected. All analyses conducted are presented and available at https://github.com/covid19climate/COVID-19-Climate.

### Statistical Analysis

Continuous variables are reported as mean ± SEM. Group differences in continuous variables were tested using the Student t-test. Correlation between weather parameters was conducted using Pearson and Spearman correlation were calculated according to data distribution. A value of P<0.05 was considered statistically significant. Statistical analysis was conducted using GraphPad Prism 6 and R studio gplot2 package.

## Results

Based on the inclusion criteria of the study, 13 countries (including Italy) fulfilled the criteria for “cold” climate countries and 7 countries fulfilled the criteria for “warm” climate countries. Four countries did not meet the “cold” nor “warm” criteria (Table 1). Confirmed COVID-19 cases according to country climate classification demonstrated a slower dynamics of case accumulation in “warm” countries. The data presented was analyzed according to the data available via CSSE as of March 19, 2020. The cumulative number of cases in a logarithmic scale is shown in Figure 1 and Supplementary Figure 1. In order to quantify the disease spread, the replication rate (the slope of the logarithmic graph smoothened for a period of 3 days) was calculated for each country (Figure 2) as well as the rate of spread the slope of the linear regression calculated for a period of 7 days (Figure 3). Mean replication rate (RR averaged over all time windows available for each country) was significantly lower in “warm” climate countries (0.12 ± 0.02) compared with “cold” countries (0.24 ± 0.01), (p<0.0001). The COVID-19 rate of spread (the slope of the linear regression of the logarithmic graph of cumulative cases) was significantly lower in “warm” climate countries (0.12 ± 0.02) compared with “cold” climate countries (0.25 ± 0.01). This difference in replication rate translates into a 2.8 times slower estimated doubling time in “warm” climate countries (7.26±1.56 days) compared with “cold” climate countries (2.89±0.16), P=0.046.

**Table 1.** Climate data, Replication rate and rate of Spread in studied countries

| Country | Climate Category | Replication rate (Mean±SEM) | Rate of Spread (Mean±SEM) | Temperature (°C) | Precipitation (mm) | Morning Humidity (%) | Evening Humidity (%) | Dew point (°C) | Wind (km/hr) |
| --- | --- | --- | --- | --- | --- | --- | --- | --- | --- |
| United Kingdom | Cold | 0.25±0.02 | 0.25±0.01 | 6.9 | 60 | 91 | 68 | 2 | 14.4 |
| Switzerland | Cold | 0.27±0.02 | 0.27±0.01 | 3.7 | 78.5 | 84 | 59 | 0 | 9.4 |
| Netherlands | Cold | 0.26±0.03 | 0.24±0.02 | 6 | 80 | 92 | 74 | 3 | 27 |
| Belgium | Cold | 0.23±0.03 | 0.22±0.01 | 6 | 80 | 88 | 68 | 2 | 22 |
| Iceland | Cold | 0.15±0.01 | 0.16±0.01 | 0.8 | 70 | 79 | 74 | -3 | 20 |
| Italy | Cold | 0.23±0.02 | 0.23±0.02 | 10.6 | 57.5 | 74 | 58 | -8 | 19 |
| Canada | Cold | 0.21±0.02 | 0.2±0.01 | -2.2 | 39.1 | 80.5 | 62.1 | 0 | 14.4 |
| Austria | Cold | 0.28±0.01 | 0.3±0.01 | 6.4 | 47.6 | 80 | 65 | 2 | 17 |
| France | Cold | 0.26±0.02 | 0.27±0.01 | 8.8 | 36 | 88 | 64 | 0 | 20 |
| Germany | Cold | 0.28±0.02 | 0.28±0.01 | 4 | 60 | 86 | 65 | -3 | 12 |
| Norway | Cold | 0.24±0.03 | 0.26±0.03 | 0.3 | 30 | 83 | 47 | 2 | 16 |
| Spain | Cold | 0.31±0.02 | 0.33±0.01 | 9.8 | 26 | 91 | 69 | -3 | 16 |
| Sweden | Cold | 0.23±0.03 | 0.24±0.03 | 0.5 | 0 | 72 | 37 | 6 | 12 |
| Egypt | Warm | 0.15±0.02 | 0.16±0.02 | 17 | 10 | 75 | 58 | 13 | 25 |
| Bahrain | Warm | 0.1±0.02 | 0.11±0.01 | 21 | 174 | 92 | 72 | 23 | 14 |
| Singapore | Warm | 0.06±0.01 | 0.05±0 | 28 | 230 | 96 | 68 | 23 | 6 |
| Malaysia | Warm | 0.2±0.03 | 0.19±0.02 | 27 | 130 | 85 | 67 | 15 | 22 |
| Australia | Warm | 0.18±0.01 | 0.17±0.01 | 17 | 30 | 88 | 55 | 23 | 9 |
| Thailand | Warm | 0.05±0.01 | 0.04±0.01 | 29 | 20 | 73 | 34 | 5 | 16 |
| Iraq | Warm | 0.11±0.01 | 0.11±0.01 | 16 | 20 | 73 | 34 | 5 | 16 |
| USA | None | 0.23±0.02 | 0.24±0.02 | 8.2 | 88.9 | 70 | 47 | -0.3 | 17.1 |
| Japan | None | 0.09±0.01 | 0.93±0.01 | 12.3 | 39.7 | 76 | 60 | 5 | 7.3 |
| Greece | None | 0.19±0.03 | 0.20±0.02 | 9.4 | 100 | 69 | 55 | 1 | 19 |
| South Korea | None | 0.19±0.04 | 0.18±0.04 | 5 |  | 74 | 47 | -2 | 12 |

**Figure 1:**
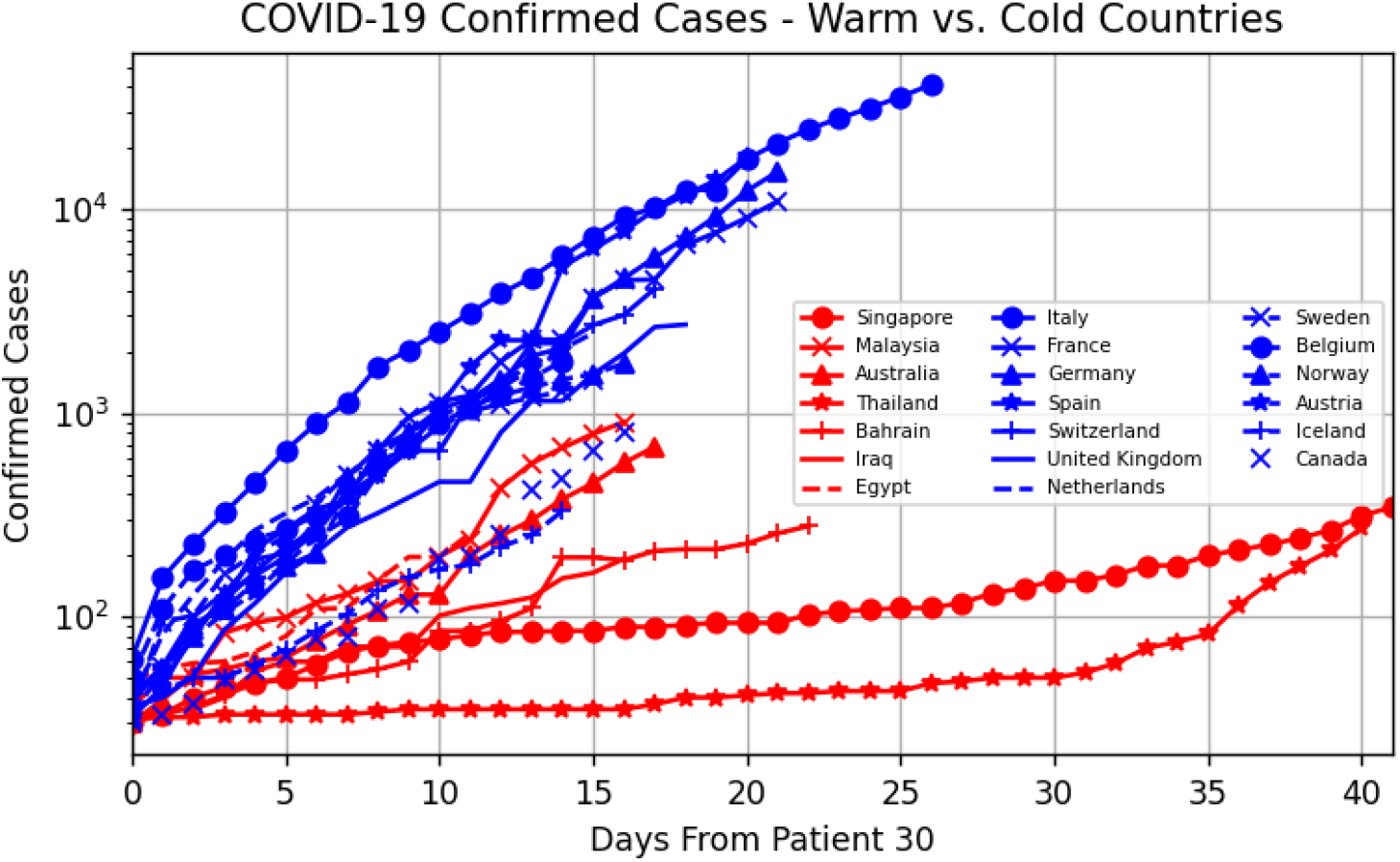
Cumulative number of COVID-19 cases in “warm” vs. “cold” countries (logarithmic scale)

**Figure 2:**
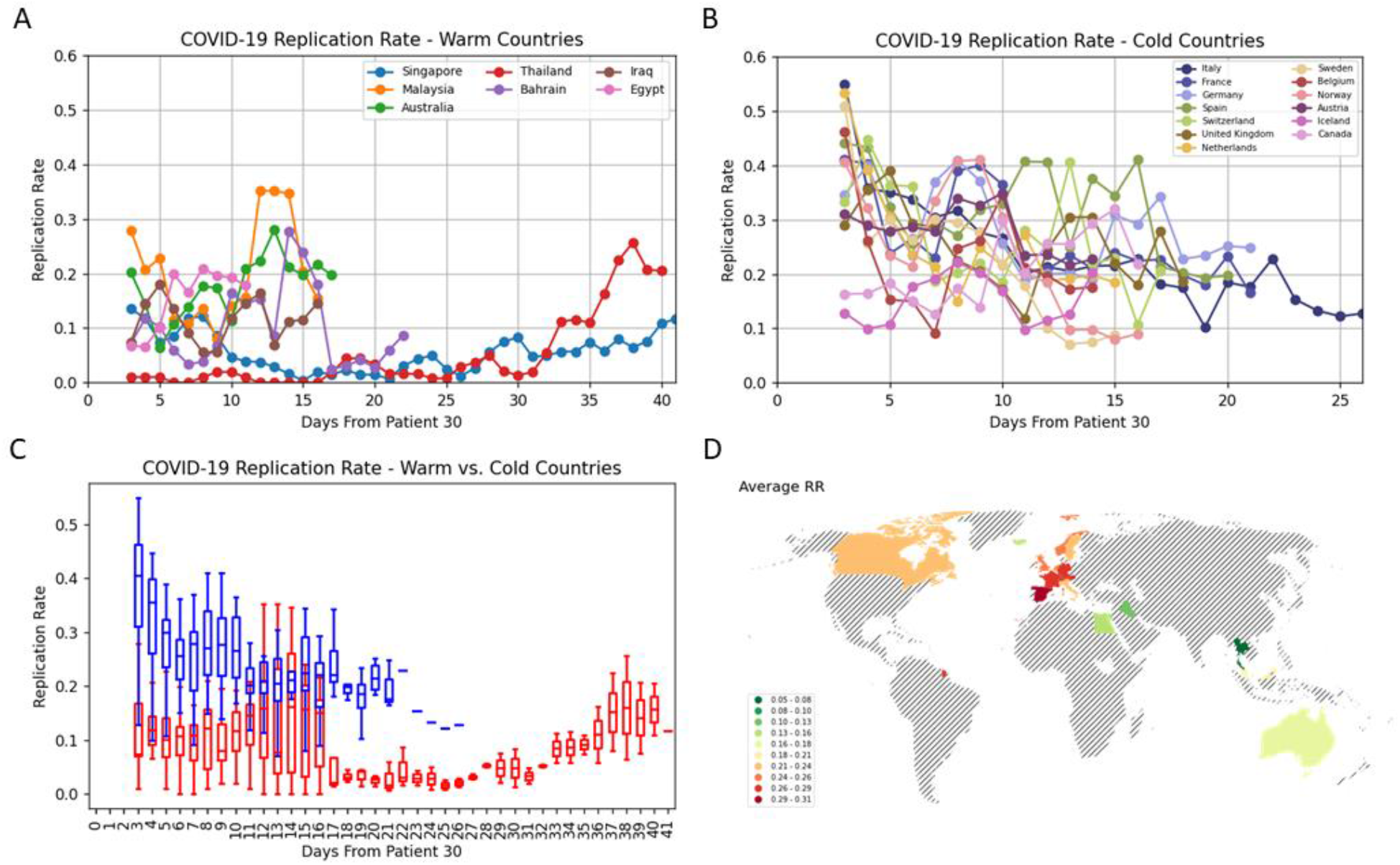
COVID-19 replication rates by country; A – “warm” countries, B- “cold” countries, C- “warm” vs. “cold” boxplot D – average replication rate on a world map

**Figure 3:**
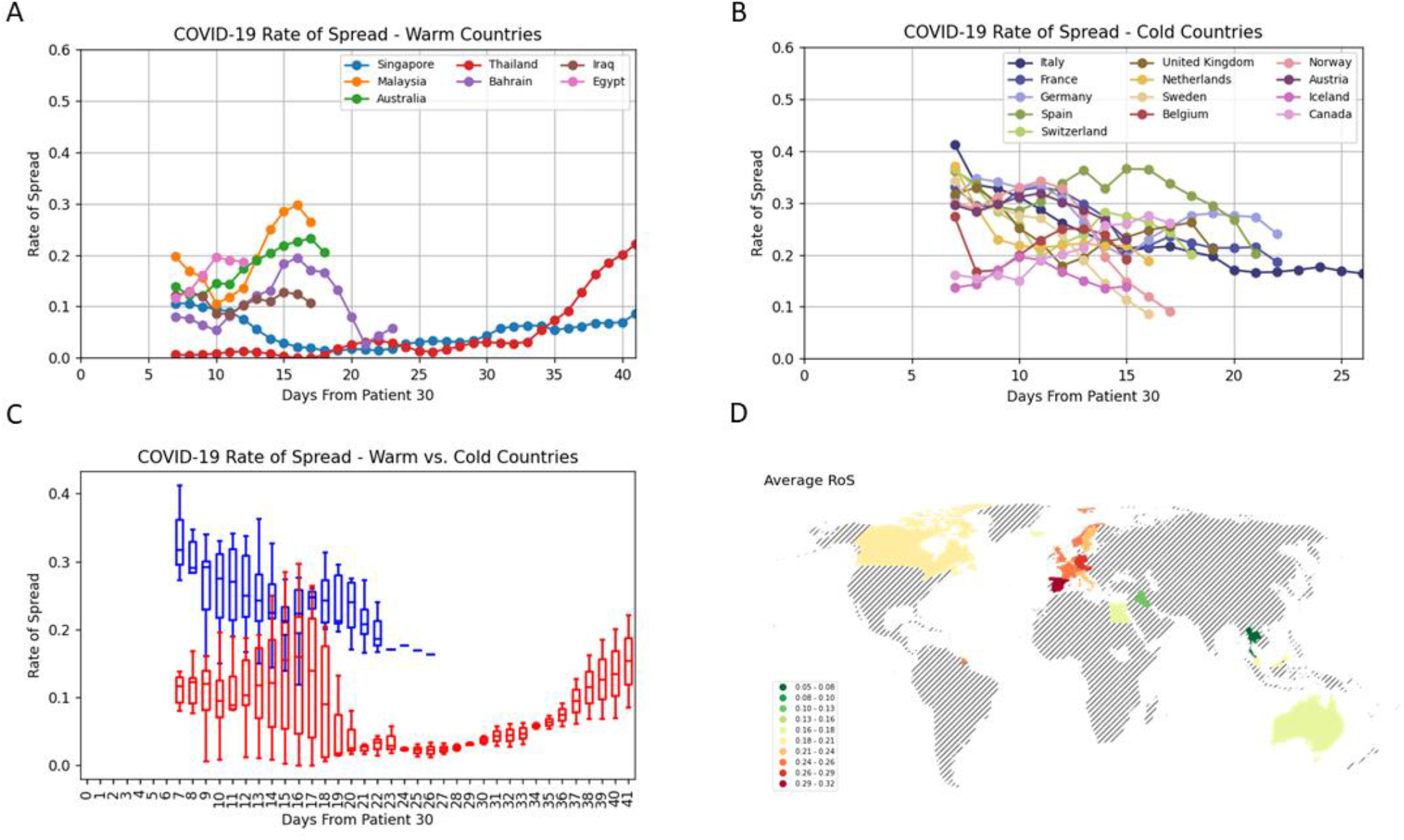
COVID-19 rate of spread by country; A – “warm” countries, B- “cold” countries, C- “warm” vs. “cold” boxplot, D- average rate of spread on a world map

We used correlation analysis in order to describe the relation between climate parameters and the RR (Table 1) and RoS (Table 2) in a non-dichotomized data set (without separation to warm and cold climate countries). Both RR and RoS displayed a moderate negative correlation with temperature R= -0.69, 95% confidence interval [CI], -0.87 to -0.36; P<0.001 and R= -0.72, 95% confidence interval [CI], -0.87 to -0.36; P<0.001, respectively (Figures 4A and Figure 5A). Climate variables such as precipitation, morning and evening humidity, and wind speed did not display a significant correlation with both RR and RoS (Figure 4 and 5). In addition, Both RR and RoS displayed a moderate negative correlation with the dew point R= -0.56, 95% confidence interval [CI], -0.82 to -0.10; P=0.02 and R= -0.62, 95% confidence interval [CI], -0.85 to -0.19; P=0.008, respectively (Figure 4C and 5C). Given the robust outbreak in Italy, most probably due to non-climate factors, we examined the discussed climate-RR/RoS correlations without the data of Italy. The correlation of RR with temperature and dew point in the dataset excluding Italy was R=-0.69, 95% confidence interval [CI], -0.87 to -0.35; P=0.001 and R=-0.59, 95% confidence interval [CI], -0.84 to -0.17 (P=0.02). The correlations of RoS to temperature and dew point remained significant as well. Therefore, we conclude that the correlations described above were unaffected by the inclusion or exclusion of Italy in the analysis. Furthermore, to exclude an effect of our pre-selected climate classification to “warm” and “cold” on the correlation between RR/RoS to climate parameters we also examined the correlation after we added countries that fulfilled our non-climate dependent criteria (USA, Japan, Greece and South Korea). Both RR and RoS significantly correlated with temperature (RR: R=-0.64, P<0.001; ROS: R=-0.67, P<0.001) and dew point (RR: R=-0.60, P=0.002; RoS: R=-0.63, P=0.001) and did not correlate with other climate parameters (similar to the correlation pattern described above without these additional countries).

**Table 2.** Correlation between replication rate and climate parameters.

| Weather Parameter | R | 95% confidence interval | p value |
| --- | --- | --- | --- |
| Temp (C°) | -0.69 | -0.87 to -0.36 | <0.001 |
| Precipitation (mm) | 0.02 | -0.44 to -0.47 | 0.95 |
| Morning Humidity (%) | 0.25 | -0.22 to 0.63 | 0.29 |
| Evening Humidity (%) | 0.12 | -0.33 to 0.53 | 0.60 |
| Dew point (C°) | -0.56 | -0.82 to -0.10 | 0.02 |
| Wind (km/h) | 0.09 | -0.37 to 0.51 | 0.70 |

**Table 3.**
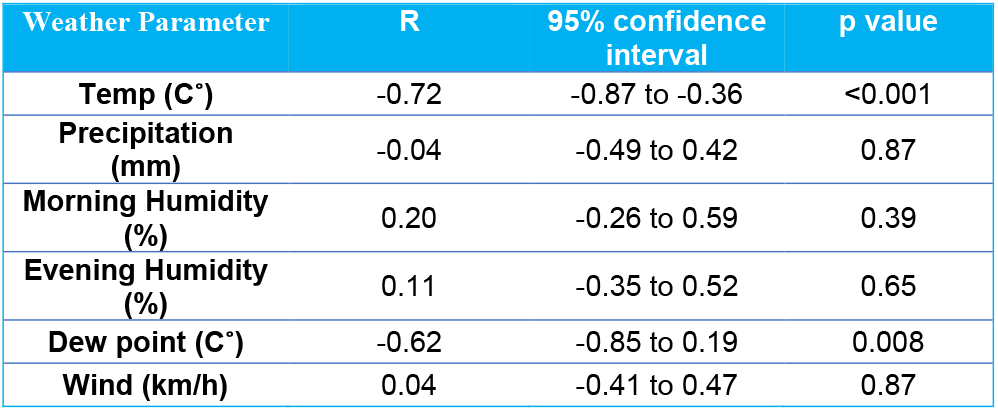
Correlation between rate of spread and climate parameters.

**Figure 4:**
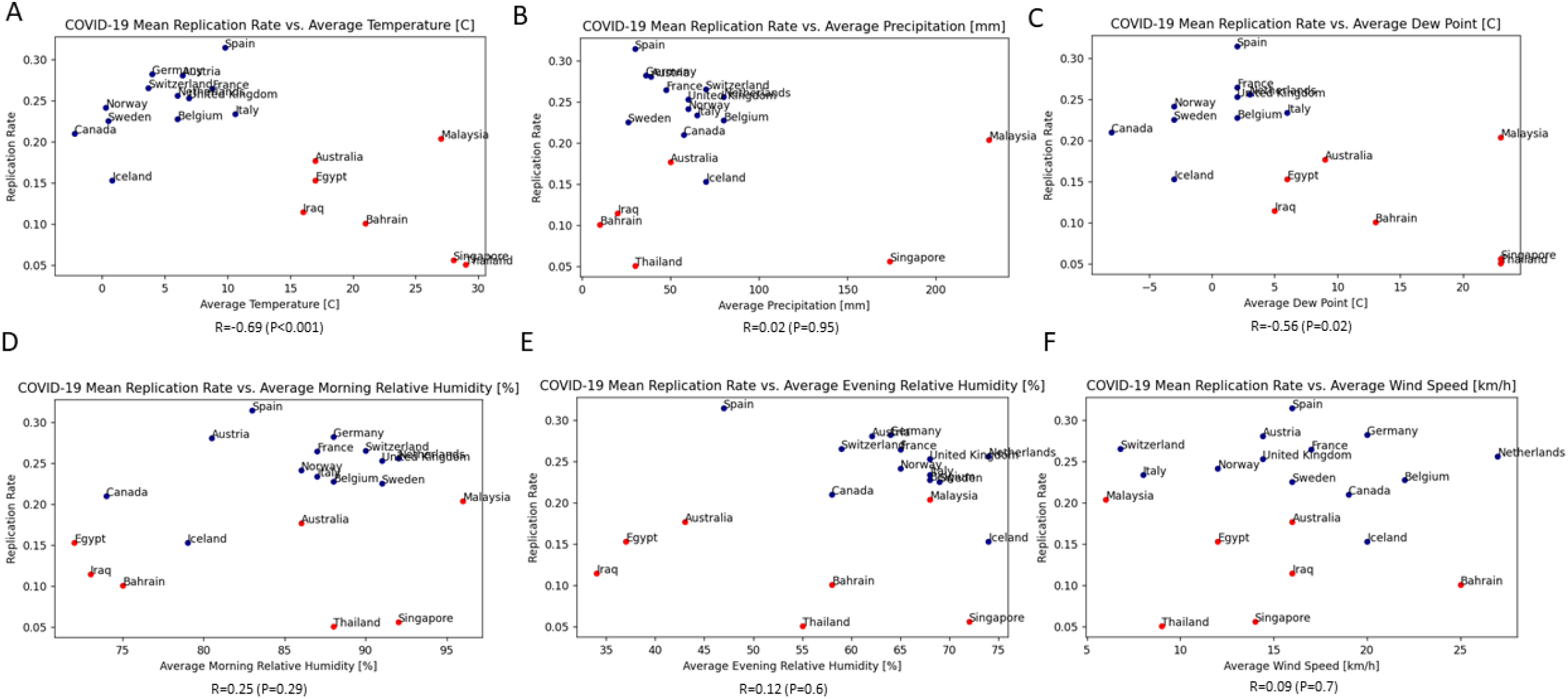
COVID-19 country replication rate and correlation to average climate parameters

**Figure 5:**
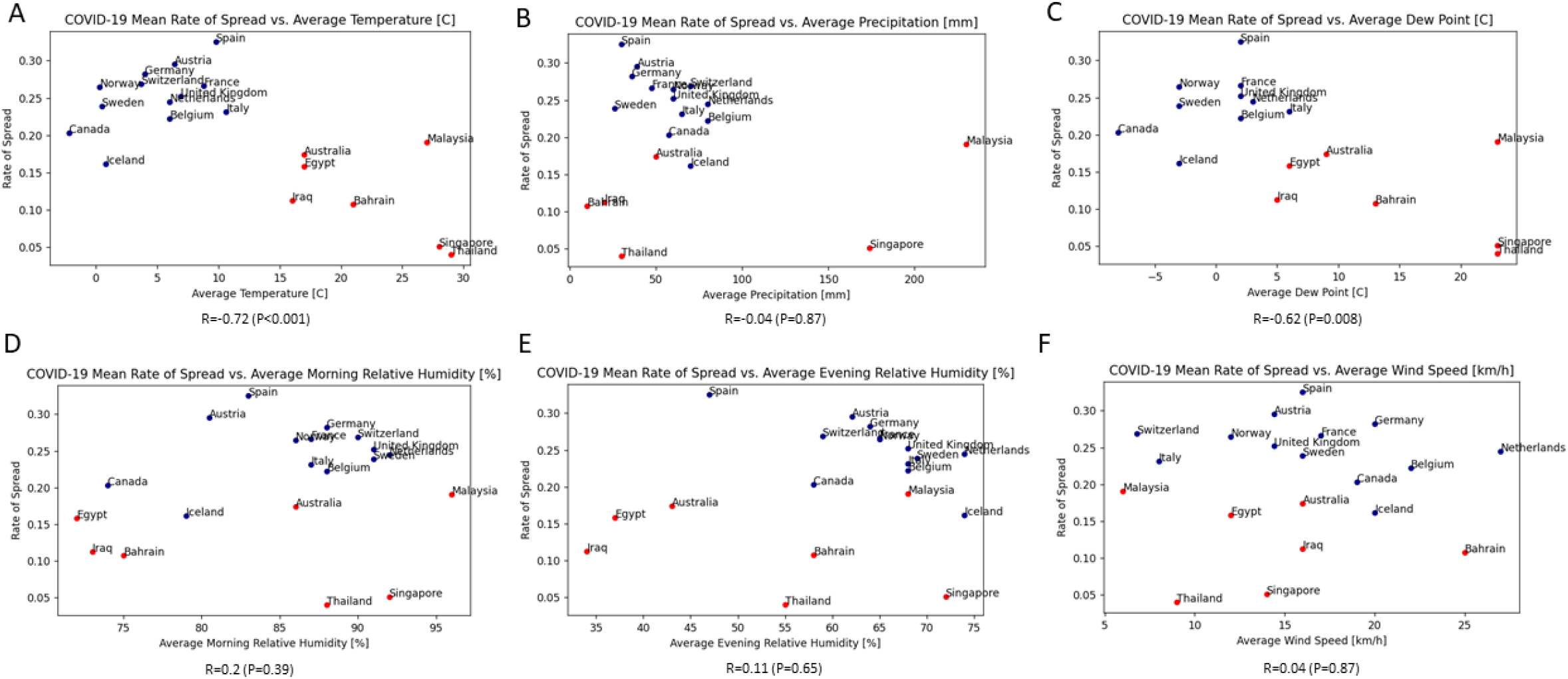
COVID-19 country rate of spread and correlation to average climate parameters

To examine the correlation with alternative parameters that might affect the RR or RoS we tested the correlation between country population size and the degree of disease spread (Supplementary Table 1). Both RR and RoS did not correlate with country population size. In addition, there was no significant difference between country population size in “cold” climate and “warm” climate countries. Finally, the number of COVID-19 tests taken may affect disease detection and thus may confound RR and RoS. However, we did not find a significant correlation between COVID-19 RR and RoS and the number of tests or the number of tests per 1000 persons. Neither was the number of tests taken significantly different between “cold” and “warm” climate countries.

Finally, we conducted an additional analysis, of intra-country correlation, within the USA between climate parameters and RR/RoS. Given the lag of disease spread in the USA, as of March, 19,2020 the cumulative patient number was limited. The RR and RoS for the USA are calculated based on limited data and this analysis was conducted aiming at future tracking on climate outbreak dynamics in the USA using the study’s dashboard as the number of cumulative cases is dynamically evolving (Supplementary Table 2). As of March 19, 2020, we did not find significant correlations between RR and RoS with any of the climate parameters evaluated (Supplementary Table 3 and 4).

## Discussion

In this study we analyzed the spread rate of the novel COVID-19 pandemic in relation to climate variables. We identified a significant moderate negative correlation between the rate of COVID-19 RR and RoS and increasing temperature and dew point. Furthermore, by dichotomizing countries into “warm” and “cold” climate (based on predefined average March temperature </>15 degrees Celsius and latitude) categories we identified a significantly lower RR and RoS in warm climate countries compared with cold climate countries. These findings persisted irrespective of whether Italy was included in our analysis or not, whether we used country dichotomization by climate criteria or included all countries irrespective of climate classification. Intra-State analysis of American (USA) data did not identify a significant correlation Our results were devised by developing an online surveillance tool (available at https://covid19.net.technion.ac.il/) coupling continuously updated COVID 19 data (available at the CCSE website^20^) with climate variables. We hope this tool will aid healthcare providers and policymakers in dynamically tracking the disease and tailoring the mitigation steps designed to slow down COVID-19 spread.

Knowledge on COVID-19 RR and RoS is sparse and interpretation is complex as it can be affected by a multitude of regional/national factors including amongst others, age distribution, cultural habits, testing & screening strategies, applied mitigation measures as well as local policy regarding administered care^21^. We found that the climate variable beside temperature, with the strongest correlation to RR and RoS was dew point, whereas precipitation, humidity and wind-speed did not appear to be significantly related to RR and RoS (Figure 4 and 5). Dew point temperature was previously shown to strongly associate with respiratory viruses spread rate and was recently found to explain approximately one third of the variation in transmission of enteroviruses across USA^22,23^. The dew point depends on both temperature and humidity and it is defined as the temperature to which air must be cooled at constant pressure for saturation to occur. Dew point may influence evaporation of aerosols containing COVID-19. Doubling time in our assessment was only marginally different between warm and cold countries, due to the high variance in RoS in warm countries.

Animal studies have suggested much lower transmission rates of influenza virus in high humidity and/or high temperature conditions^24^.Our findings of lower RR and RoS in warm climate may suggest that the seasonal climate variation will influence disease spread dynamics globally in the coming months. Colder temperatures may provide better conditions for virus survival outside the human body, with longer viral viability on contaminated surfaces and fomites. The SARS-CoV survives longer in colder and less humid conditions on contaminated surfaces^25^. In preliminary findings, Bannister Tyrrell et al have demonstrated that higher average temperature was strongly associated with lower COVID-19 incidence^26^.

We developed this interactive online surveillance tool that tracks the correlation between the global climate parameters and COVID-19 as a service to leadership, healthcare providers and the general public. It is readily reproducible, updates virtually online and enables examination of the correlation between the various climate variables and COVID-19 replication rate over time. The code was written using the Jupyter notebook environment via Google Colaboratory enabling researchers from countries with lesser available computing resources to run and execute their version of the code freely in the cloud. We hope it will serve healthcare organizations and leadership in deciding which mitigation steps to further take while also accounting for climatic alterations.

The COVID-19 outbreak occurred during the winter season mostly in the northern hemisphere and the correlation of outbreak spread rate and climate is continuously studied. A recent study pinpointed to the observation that several regions with COVID-19 outbreaks occurred within the same latitude range, in areas with low temperatures and high humidity as of March 5, 2020^10^. A preliminary clue for a possible association between temperature and COVID-19 was reported online from Bannister-Tyrrell et al.^27^ Our results in the current study may strengthen the rationale to further assess the association between climate parameters and COVID-19 outbreak spread rate. Given the preliminary association identified in our study as well as by others, between the latter parameters, we advocate for prospective surveillance of disease trajectories in relation to contemporary climate data.

There are several strengths in our findings. Our data is updated through March 19 2020, by which many countries imposed international restrictions, ensuring that our findings are more representative of local spread rather than imported cases. Moreover, we chose to look at data beyond patient 30 (for countries and USA states), further ensuring that the diagnosed cases are from local spread. We demonstrated that average temperature as well as dew point, were negatively correlated with the RR and RoS in the worldwide analysis. Twenty four countries were included in our analysis which aids in generalizing these findings worldwide as the pandemic rages on. In addition, we demonstrated that potential confounders such as the population size, the number of tests and the number of tests/1000 persons did not significantly correlate with RR or RoS. Finally, our results are easily reproducible and the developed tool will help track the dynamic changes and reassess the correlation between climate and disease spread in the future.

The current study has several limitations. Different mitigation steps were undertaken by each included country during different times affecting the spread of COVID 19 in ways we cannot account for. However, since these mitigation steps have a lag period of approximately14 days (based on the virus incubation time) they may only marginally affect our analysis. Also, as most of our “cold” climate countries are located in Europe, it important to note that ground travel within the European Union was unrestricted until very recently, a variable which may increase the imported spread rate. The lack of standardized criteria for diagnostic testing for COVID-19 between countries affects the incidence and cumulative count. Be that as it may, number of tests performed in each country did not correlate with the RR and RoS. We used the climate parameters of the capital city of each country (and state in the USA) to represent each country in its entirety in order to calculate the RR and RoS as more detailed locations of COVID-19 diagnoses in each country were not available to us, thus local spread within the country was not accounted for. An additional limitation that may warrant further research is the reliance on historical average temperatures. Further research and update of the surveillance tool will allow tracing these dynamic trends in relation with real-time temperature and the appropriate time lag consistent with COVID-19 incubation period. Finally, our results might further be confounded by the varied socioeconomic status of the participating countries as well as social context.

COVID-19 RR and RoS are affected by a multitude of factors. As the pandemic is spreading across the world with an alarmingly increasing toll of diagnosed cases as well as deaths, our findings of decreased RR and RoS in warm climates may suggest that the inevitable seasonal variance will alter the dynamic of the disease spread in both hemispheres in the coming months. However, we warrant a cautious interpretation of these findings given the fact that we are in the initial steps of this outbreak in many “warm” climate countries, the high variance of the data and the dynamic changes in the disease surveillance and the lack of correlation based on the limited data in the US. The current evidence from our study, does not justify any modification of governmental mitigation strategies in countries with warm climates. We hope that the online tool coupling COVID-19 data with climate data will assist in tracking the disease and tailoring the needed measures to contain it.

## Data Availability

Data will be available upon request.

https://covid19.net.technion.ac.il/

## Figure Legends

**Figure 1**. Cumulative COVID-19 confirmed cases (logarithmic scale) of “warm” (red) and “cold” (blue) countries in days from patient 30.

**Figure 2**. Replication rate of “warm” countries (A), “cold” countries (B), “warm” vs. “cold” countries boxplot showing the median line, a box between quartile 1 and quartile 3 (Q1-Q3) and whisker at the size of 1.5x(IQR-Interquartile range) (C) in days from patient 30. Average replication rate in “warm” and “cold” countries on a world map (D). RR-Replication Rate.

**Figure 3**. Rate of spread of “warm” countries (A), “cold” countries (B), “warm” vs. “cold” countries boxplot showing the median line, a box between quartile 1 and quartile 3 (Q1-Q3) and whisker at the size of 1.5x(IQR-Interquartile range) (C) in days from patient 30. Average rate of spread in “warm” and “cold” countries on a world map (D). RoS-Rate of Spread.

**Figure 4**. Scatter plot representation of replication rate correlation to climate parameters in “warm” (red) and “cold” (blue) countries. A - average temperature [°C], B - average precipitation [mm], C - average dew point [°C], D - average morning relative humidity [%], E – average evening relative humidity [%], F – wind speed [km/h].

**Figure 5**. Scatter plot representation of rate of spread correlation to climate parameters in “warm” (red) and “cold” (blue) countries. A - average temperature [°C], B - average precipitation [mm], C - average dew point [°C], D - average morning relative humidity [%], E – average evening relative humidity [%], F – wind speed [km/h].

**Supplementary Figure 1**. Cumulative COVID-19 confirmed cases (logarithmic scale) of “warm” (A) and “cold” (B) countries in days from patient 30.

**Supplementary Figure 2**. Doubling time in “warm” and “cold” countries on a world map.

**Supp. Figure 1:**
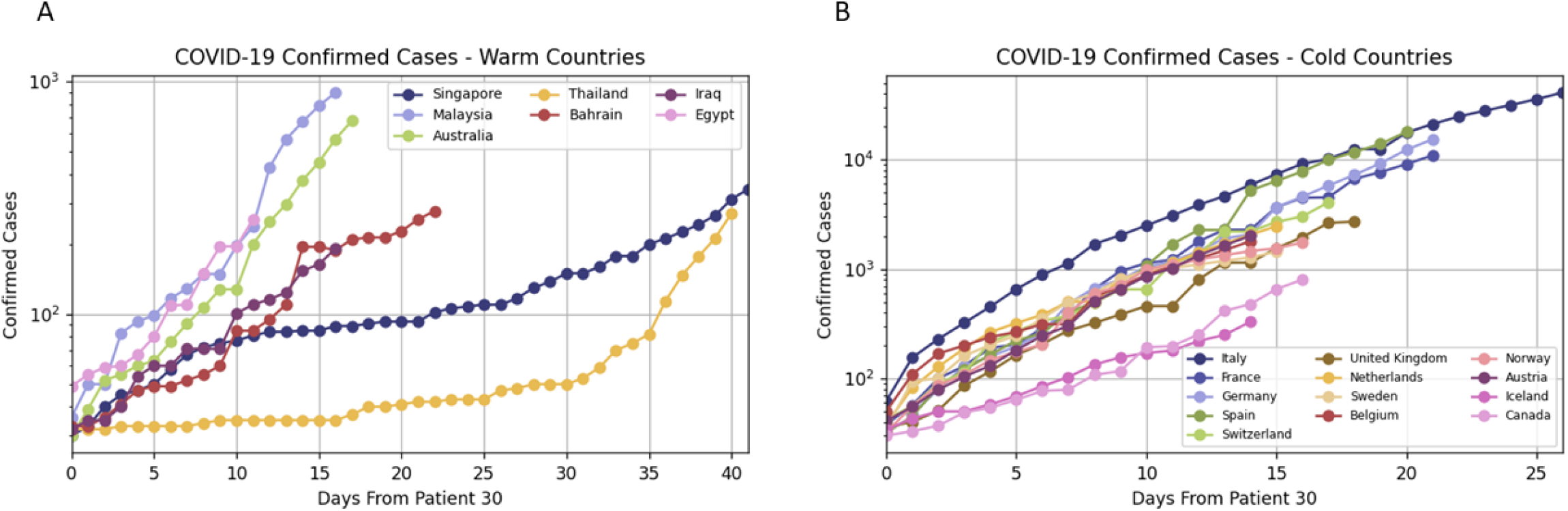
Cumulative number of COVID-19 cases in “warm” and “cold” countries (logarithmic scale); A- cold countries, B- warm countries

**Supp. Figure 2:**
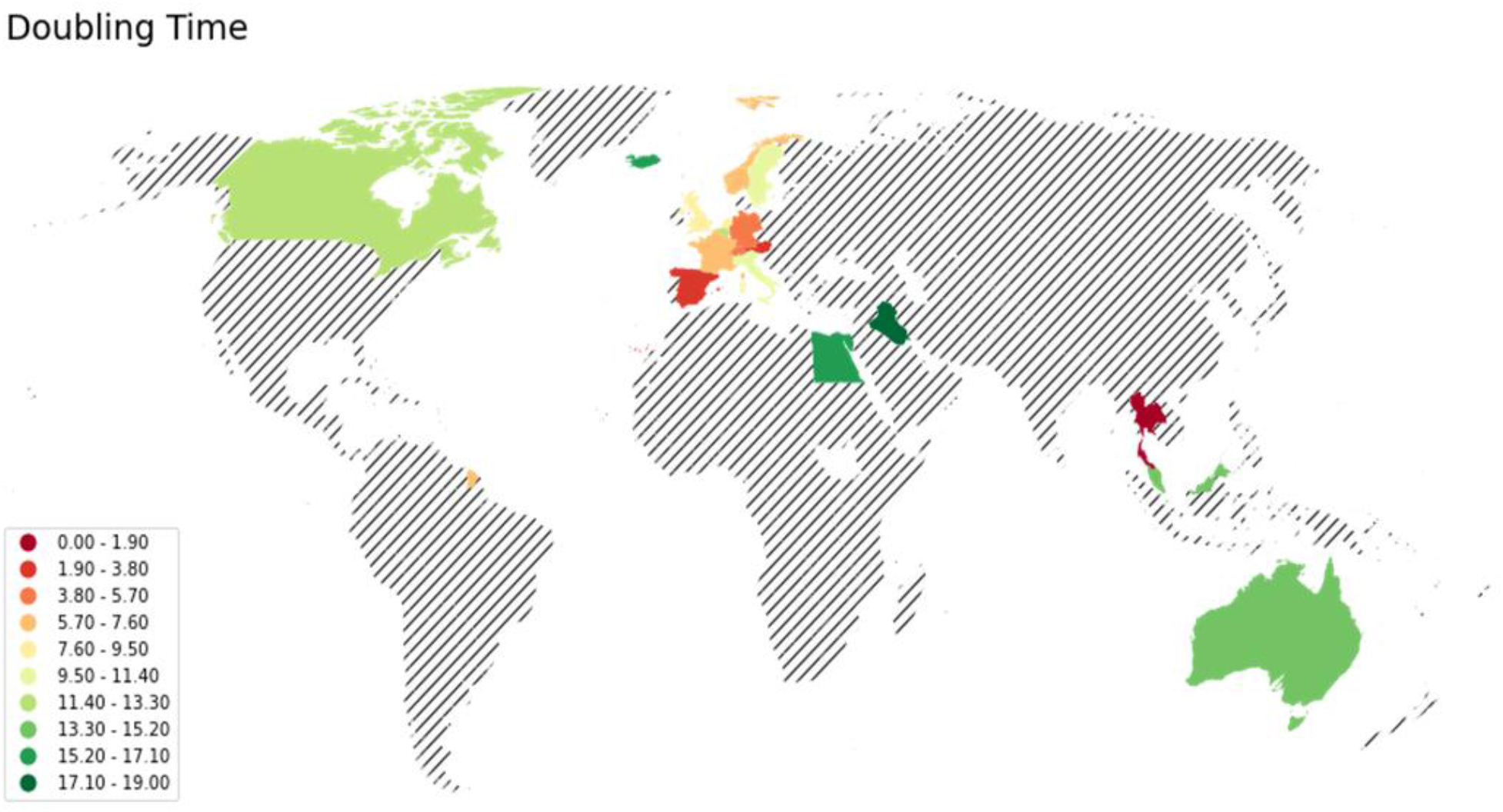
Doublingtime in “warm” and “cold” countries on a world map

